# Recovery of kicking kinematics and performance following intermittent high-intensity running bouts in young soccer players: can a local cooling intervention help?

**DOI:** 10.1101/2022.03.05.22271957

**Authors:** Luiz H Palucci Vieira, Christopher Carling, Carlos A Kalva-Filho, Felipe B Santinelli, Lorenzo A G Velluto, João Pedro da Silva, Eleftherios Kellis, Fabio A Barbieri

## Abstract

Repeated high-intensity running (RHIR) exercise is known to affect central and peripheral functioning. Declines in RHIR performance are exacerbated by environmental heat stress. Accordingly, the use of post-exercise cooling strategies (COOL) is recommended as it may assist recovery. The present study aimed to investigate, in a hot environment (> 30ºC), the effects of local COOL following RHIR on indices of soccer kicking movement and performance in youth soccer. Fifteen academy under-17 players (16.27 ± 0.86 years-old; all post-PHV), acting as their own controls, participated. In #Experiment 1, players completed an all-out RHIR protocol (10 × 30 m bouts interspersed with 30 s intervals). In #Experiment 2, the same players performed the same running protocol under two conditions, 1) 5 minutes of COOL where ice packs were applied to the quadriceps and hamstrings regions and, 2) a control condition involving only passive resting. In both experiments, perceptual measures [ratings of perceived exertion (RPE), pain and recovery], thigh temperature and kick-derived video kinematics (hip, knee, ankle and foot) and performance (ball speed and placement) were collected at baseline and post exercise and intervention. In the first experiment, RHIR led to moderate-to-large increases (*p* < 0.03) in RPE (*d* = 4.08), ankle eversion/inversion angle (*d* = 0.78) and mean radial error (*d* = 1.50) and small-to-large decreases (*p* < 0.04) in recovery (*d* = -1.83) and average/peak ball speeds (*d* = -0.42–-0.36). In the second experiment RPE (*p* < 0.01; Kendall’s *W* = 0.30) and mean radial error (*p* = 0.057; η^2^ = 0.234) increased only post-control. Significant small declines in ball speed were also observed only post-control (*p* < 0.05; *d* = 0.35). Post-intervention CM_foot_ velocity was moderately faster in COOL as compared to control (*p* = 0.04; *d* = 0.60). RHIR acutely impaired kicking movement, ball speed and placement in youth soccer players. However, a short period of local cryotherapy may be beneficial in counteracting declines in indices of kicking performance in hot environment.

**Trial registration number:** #RBR-8prx2m - ReBEC Brazilian Clinical Trials Registry

## INTRODUCTION

In young soccer players, accumulated fatigue is manifestly observed over the course of matches. This is illustrated by an increase in players’ ratings of perceived exertion (RPE) and a concomitant reduction in running outputs during the second half (Aslan et al., 2012). Although the majority of in-game activities (∼75–82%) are performed in the ‘low intensity’ domain (< 13 km/h; Buchheit et al. (2010a)), the repeated execution of high-intensity running bouts (Buchheit et al., 2010b) is associated with substantial acute increases in both perceived effort (Brocherie et al., 2015; Sánchez-Sánchez et al., 2014) and sensations of pain (Monks et al., 2017). These can be linked to impairments in neuromechanical responses, explained by either central or peripheral fatigue factors (Brocherie et al., 2015; Goodall et al., 2015; Perrey et al., 2010). In studies using protocols to simulate the running loads commonly found in official matches, including intermittent high-intensity activities, also identified declines in indices of kicking performance (Palucci Vieira et al., 2021b; Russell et al., 2011; Sánchez-Sánchez et al., 2014), notably in senior players. In contrast, there is only limited evidence level regarding the magnitude of the effects of repeated high-intensity exercise on kicking performance in youth populations [for a review: (Palucci Vieira et al., 2021b)].

In an attempt to accelerate recovery processes during/following exercise, cooling interventions are frequently utilised and their beneficial effects are seemingly amplified in hot environments (Bongers et al., 2015). For example, following exercise, cooling may have a transient analgesic effect aiding reduction of swelling and muscle pain and lowering RPE values (Bleakley et al., 2012; Duffield et al., 2013) thus facilitating the execution of subsequent movement/physical effort (Fischer et al., 2009). Some studies have demonstrated short-to-long-term beneficial effects of cooling on running outputs in soccer players notably post-match–periods of fixture congestion–or during the half-time interval (Buchheit et al., 2011; Duffield et al., 2013). However, the literature generally shows that cooling techniques can exert acute negative effects on motor performance immediately after an intervention. These effects include severe declines in lower limb power-dependent activities (e.g. jumping and running at maximal speed) and goal-directed technical skills (Bleakley et al., 2012; Tyler et al., 2015; Wassinger et al., 2007). Longer cooling treatment times utilised in the majority of interventions (e.g. up to 20 minutes) can notably be an issue (Bleakley et al., 2012). As such, a shift to using shorter cryotherapy durations is arguably necessary (Egaña et al., 2019; Fischer et al., 2009; Peiffer et al., 2010) as well as having practical implications. Notably, preliminary evidence in both individual and team sport athletes has shown that rapid post-exercise cooling (≤ 5 minutes) may reduce the negative effects of exercise on power development (Egaña et al., 2019; Peiffer et al., 2010) or at least not impair peripheral blood flow, muscle temperature and motor responses (Fischer et al., 2009; Thorsson et al., 1985; Zemke et al., 1998).

It is clear from the above discussion that recovery-related strategies are necessary in effectively re-establishing explosive performance following intense physical exercise demands (e.g. repeated high-intensity running). Yet, the potential benefits of recovery methods including cooling techniques that are ubiquitously used in soccer contexts on kicking parameters are still unknown (Palucci Vieira et al., 2021b). Local application of ice packs has previously demonstrated positive effects in reducing perceived pain (Algafly and George, 2007), improving thermal and recovery sensations (Wiewelhove et al., 2020) and even assisting the power response of lower limbs in a heat stress experimental condition (Castle et al., 2006). The rolling substitute policy used in youth soccer tournaments played in hot climatic conditions could notably benefit from this mode of cooling to help alleviate game-related load demands in substitute players on pitch entry. To the extent of our knowledge however, the lack of information related to the effects of recovery treatments such as cooling strategies is even greater in relation to soccer technical skills. A recent systematic review has identified a lack of studies investigating the influence of cooling specifically in goal-directed skills, particularly in soccer kicking performance, and notably where concomitant demands for precision and velocity are required (Palucci Vieira et al., 2021b). Thus, the aim of the present study was to verify, under conditions of thermal (heat) stress (temperature > 30ºC; Girard et al. (2015)), the effects of local cooling following intermittent high-intensity efforts on kicking movement kinematics and performance in young soccer players. We hypothesized that an all-out running exercise in the heat would generate acute reductions in ball kicking movement and outcomes, and notably velocity outputs. We then expected that the subsequent application of ice pack during a short period (5 minutes) would favour youth soccer players perceived well-being (e.g. sensations of recovery, pain and exertion) while not negatively interfering with kick movement or performance recovery after intense exercise in the heat.

## MATERIALS AND METHODS

### Participants

Fifteen youth players participated (16.27 ± 0.86 years-old; 2.12 ± 0.71 years from peak height velocity; 64.14 ± 10.98 kg; 172 ± 9 cm). All procedures were approved by the local Human Research Ethics Committee (protocol #2650204; CAAE85994318.3.0000.5398) and Brazilian Clinical Trials Registry ReBEC (http://www.ensaiosclinicos.gov.br/; included in the network of WHO primary registries) under number RBR-8prx2m. In #Experiment 1 which tested exercise-induced changes in kicking parameters, a subsample of 13 players was evaluated. This was considered to be the required sample size estimated owing to the expected declines in kicking performance following general intermittent high-intensity exercise mode (effect size = 0.92; power = 85% and α = 0.05) [data from a systematic review by Palucci Vieira et al. (2021b)]. In #Experiment 2 which evaluated the effects of the recovery intervention, all 15 players were evaluated to meet the a priori required sample size based on the assumption that ice would affect motor skills precision (effect size = 0.86; power = 80% and α = 0.05) (Wassinger et al., 2007) and/or global performance in immediately subsequent sports-related tasks (average effect size = 0.83; data from systematic review by Bleakley et al. (2012)). Sample size estimations for each experiment were obtained using G*Power© v.3.1.9.2 environment (Universität Düsseldorf, Germany). The players were invited from the under-17 age-group of a club that competes at state standard in Brazil (1^st^ place in São Paulo Interior League 2020 edition). Both youth athletes and their legal guardians signed respectively approved assent and consent forms to allow participation.

### Experimental design

In #Experiment 1, a pre-post paired test design was used, in order to analyse the impact of the running exercise protocol on measures of kicking performance. Players were firstly asked to perform a standardised 15-minute warm-up (dynamic stretching, jogging and submaximal kicks). Thereafter, they performed a running protocol involving high-intensity intermittent exercise (see below). Immediately prior to and at end of this exercise stimuli, players undertook a kicking protocol that allowed monitoring of lower limb movement mechanics and performance.

In #Experiment 2, a repeated measures pre-post testing design was adopted, randomised and counterbalanced between data collection days, where subjects acted as their own controls. After completing the same warm-up and running protocol as in Experiment 1, each participant was assigned to one of the 2 experimental conditions [control or 5 minutes cooling (COOL)] on two separate days, 24–26 hours apart. All subjects performed the kick testing protocol before commencing the running exercise in a rested state and immediately following the post-exercise intervention with COOL or control (5 minutes of passive recovery).

Both experiments were performed on an official FIFA-standard natural grass soccer pitch in the presence of sunlight during afternoon period (between ∼14:30–17:30 h). The recorded environmental temperature and relative humidity (provided by an automatic station of Centro de Meteorologia de Bauru, Local Meteorological Research Institute IPMet–UNESP–Bauru–Brazil; https://www.ipmetradar.com.br/) in Experiment 1 were respectively 36.67 ± 3.3ºC [32–41ºC] and 26.70 ± 8.78% [15.10–35.90%]; in Experiment 2 35.5 ± 2.8ºC [33.37–38.67ºC] and 20.2 ± 7.5% [15.12–28.8%] (control condition) and 33.8 ± 4.6ºC [30.59–39.09ºC] and 21.0 ± 7% [14.82–28.55%] (COOL condition).

### Kick testing collection and data processing

The kick testing protocol adopted was based on that employed in a previous study (Palucci Vieira et al., 2022b). In brief, the participants were asked to perform instep kicks, 18 m from the midpoint goal line, using FIFA-approved stationary balls (PENALTY® brand, 5-sized, 70 cm diameter, 430 g weight, and air pressure kept at 0.7 atm). Kicks were performed at maximal velocity and aimed at the centre of a 1 × 1 m target fixated in the contralateral goalpost upper corner. The approach run was constrained to 3.5 m and 45 degrees, with a 40 s passive rest interval between repeated attempts within the same block. Differences to the original protocol included: trying to increase standardization as much as possible between the two proposed intervention conditions as well across the time-moments, no goalkeeper was used, and three kick attempts were allowed per block (time-moment).

Body motion and ball displacement immediately after kicking were recorded using two digital video cameras fixed on tripods (GoPro Hero 7 Black Edition, GoPro GmbH, München–Germany), sampling at 240 frames/s [wide field-of-view (FOV) mode; 1280 × 960 pixel; 1/480 s shutter speed], were turned on and synchronised via remote control (Smart Remote GoPro). The cameras were positioned laterally around the kick mark (2.5 m) so that their focus had ∼90 degrees between them. Afterwards, video files from data collections were downloaded onto a laptop computer (DELL INSPIRON 5590; Dell Inc., Texas–USA). The OpenPose markerless motion detector method in addition to a tracking algorithm previously validated to evaluate ball kicking action (Palucci Vieira et al., 2022c) were used to automatically extract 2-dimensional screen coordinates of seven keypoints derived from the hip (preferred and non-preferred), knee, ankle and foot regions (measurement error = 3.49 cm and 1.29 m/s; Palucci Vieira et al. (2022c)). A calibration frame was defined using 49 reference points with absolute 3-dimensional coordinates known (4.11 × 4.05 × 1.30 m). Following tracking and appropriate correction of the radial distortion (Rossi et al., 2015), screen coordinates of both cameras were inputted in a specific Python 3.8.3 algorithm (Python Software Foundation, Delaware–USA) to run 3-dimensional Direct Linear Transformation (DLT) reconstruction. Time-series positional data was then extrapolated following impact (20%) and smoothed (dual filter 4^th^-order Butterworth/rloess) in MATLAB software (R2019a MathWorks Inc., Natick–USA). Residual analysis helped define filtering parameters (cut-off frequency=25 Hz and span=0.1, respectively). After treatment, extrapolation was then removed. Custom-built routines were written to obtain linear velocities (non-preferred hip, CM_foot_ and CM_foot_ to knee relative), angular (i) joint displacement (ankle plantarflexion/dorsiflexion and eversion/inversion at impact) (ii) range-of-motion (hip and knee flexion/extension) and (ii) peak knee extension velocity. These were computed using local reference frames of joints and segments as described elsewhere (Palucci Vieira et al., 2021a; Palucci Vieira et al., 2022a).

In an attempt to compute ball speed metrics, the ball centroid was manually tracked by the DVIDEOW kinematic system (Rossi et al., 2015), using image sequences from both cameras, considering 10 available airborne frames after the foot contacted the stationary ball. To determine resultant post-impact ball speed, its horizontal and vertical components were calculated from the first derivative of linear and quadratic (second derivative = -9.81 m/s^2^) regression lines, respectively (Nunome et al., 2006). Mean and maximal values for ball speed across each block of three kicks were retained for further analysis.

To obtain ball placement-derived indices, two cameras sampling at 60 frames/s (GoPro Hero 7 Black Edition, GoPro GmbH; 1920 × 1080 pixel, linear FOV) were placed one in front of the goal (∼23 m apart) and another above the goal line. A calibration frame was defined considering all goalpost upper/lower extremities (four reference points; 7.35 × 2.32 m). The 2-dimensional coordinates of the ball centre at the moment it crossed the goal line were obtained using the same software and similar procedures as for ball speed digitisation. The Euclidean distance between the ball and target centre coordinates was calculated for each kick attempt. Taking the three repeated kick attempts within a same given block, the mean radial error (average ball-target distance), bivariate variable error (square root of the sum of standard deviation squared derived from x- and y-coordinates of the ball) and overall accuracy (a compound of the two previous measures) were computed using specific equations as described elsewhere (Vieira et al., 2018) where Euclidean distances and 2-dimensional coordinates of ball and target centre were adopted as input parameters.

### Running protocol

To simulate repeated high-intensity running efforts that players frequently undertake during soccer training and testing, a protocol including 10 “all-out” running bouts x 30 m distance each, interspersed by a recovery period of 30 s was conducted. In particular, the player ran for 25 s at low intensity back to the starting line, ensuring 4–5 s of passive resting before performing the next sprint repetition (Buchheit and Mendez-Villanueva, 2014). In young soccer players of various ages, this RHIR model has previously demonstrated good construct validity for predicting in-game running performance (Buchheit et al., 2010a). Standardised (“go, go, go …”), constant and strong verbal encouragement was provided during each effort. The time to complete each bout was recorded by a single experienced examiner using a digital manual stopwatch (LIVEUP® SPORTS, Paraná–Brazil; 1/100 s sensitivity).

### Ice application

At the end of the running exercise, the participants performed the cooling protocol (COOL condition). They were asked to sit on the substitutes bench pitch-side near to where the kicking and running protocols took place. A licensed physical therapist tightly covered the quadriceps and hamstrings of the participant’s preferred lower limb using plastic wrapping paper, respectively with two thin plastic bags (20 × 40 cm), approximately 1/3 filled with cubed ice; these were maintained constantly over muscle sites for 5 minutes (Algafly and George, 2007; Fischer et al., 2009). During the ice application, participants kept their treatment leg comfortably extended on an auxiliary chair at a height slightly lower than the bench on which they were sitting.

### Perceptual measures

Before the beginning (Pre), immediately following the running cessation (post-RHIR) and intervention or control conditions (Post), measures referring to ratings of perceived exertion were collected, using the 0–10 Borg scale (Foster, 1998); subjective perception of pain, using a 10-point Likert scale (0 = no soreness and 10 = very, very sore) (Pointon et al., 2011) and perception of recovery, based on another 10-point Likert scale (0 = very poorly recovered/extremely tired and 10 = very well recovered/highly energetic) (Paul et al., 2019). The skin surface temperature, at the midpoint of the thigh, was also determined at these same time-moments by a single examiner using an infrared manual thermometer (precision = ± 0.2ºC; capture range = 0–60ºC; model YRK-002A – HC260, Multilaser Industrial S.A., São Paulo–Brazil).

### Statistical analysis

Statistical tests were performed in IBM Statistical Package for the Social Sciences v.25 (IBM Corp. ©, Armonk–USA) with an alpha level set at *p* ≤ 0.05 for determining significance unless otherwise stated. Data normality was firstly assessed using the Shapiro-Wilk’s test. If data was flagged as non-normal, then kurtosis, skewness and frequency plots were checked. If log-transformation was not efficient, non-parametric versions of tests were used. In #Experiment 1, Student’s t test for dependent samples was used to compare measures pre- and post-RHIR. Effect size for paired comparisons was obtained using Cohen’s *d* where *d* > 0.20 (small), > 0.50 (medium), and > 0.80 (large). In #Experiment 2, Student’s t test or Wilcoxon signed-rank test was employed to obtain estimates of reliability of the responses to the running protocol between testing days/conditions. As in the case where the latter was necessary, r effect size (r = z/√N) was calculated and interpreted as r > 0.10 (small), > 0.30 (moderate) and ≥ 0.50 (large). Intraclass correlation coefficients (ICC), typical error (TE) and coefficient of variation (CV) were also computed using a specific Microsoft Excel (Microsoft Corp., Redmond–USA) spreadsheet (x.rely.xls, available on https://sportsci.org/). Finally, to compare the two distinct recovery interventions, 2 (time: pre, post) x 2 (condition: Control, COOL) repeated measures ANOVAs were run with Bonferroni adjustment to the alpha level in post-hoc comparisons. Partial eta-squared (η^2^) was taken as effect size for main effects and deemed as η^2^ > 0.01 (small), > 0.06 (moderate), and > 0.15 (large). When necessary, Friedman’s two-way ANOVA by ranks was used, also with post-hoc significance adjusted by dividing alpha level to the number of multiple comparisons performed. Kendall’s *W* effect size for main effect was determined and considered as *W* > 0.10 (small), > 0.30 (moderate) and ≥ 0.50 (large).

## RESULTS

### #Experiment 1

#### Intense running exercise, perceptual measures and kicking outputs

The repeated high-intensity running protocol led to a significant *large* increase in ratings of perceived exertion (percentage difference, mean/median absolute difference [CI lower; upper] = +659.74%, 4 a.u. [3; 9]; *p* < 0.01) and a *large* decrease in perception of recovery (−40.47%, 3.62 a.u. [1.66; 5.57]; *p* < 0.01). A *large* significant increase in mean radial error was observed (Table 1) following the exercise protocol (+34.90%, 0.67 m [0.10; 1.25]; *p* = 0.03). There was also a *large* non-significant increase in accuracy (+18.12%, 0.50 m [0.14; 1.14]; *p* = 0.11). *Small* significant declines occurred in average (−3.14%, 0.89 m/s [0.06; 1.73]; *p* = 0.04) and peak ball speed values (−3.94%, 1.18 m/s [0.15; 2.21]; *p* = 0.03). Ankle eversion/inversion angle of the kicking limb at impact *moderately* increased after the running protocol (+257.14%, 0.18 rad [0.04; 0.33]; *p* < 0.02).

**Table 1.** Effects of repeated high-intensity running bouts (RHIR) on perceptual measures and kicking performance indices (n = 13).

|  | Pre-RHIR | Post-RHIR | *p*-value | ES [90% CL] – rating |
| --- | --- | --- | --- | --- |
| RPE (a.u.) | 0.77±1.17* | 5.85±2.54 | 0.006 | 4.08 [2.96; 5.19] – large |
| Pain (a.u.) | 1.31±0.48 | 1.54±0.78 | 0.337 | 0.45 [-0.35; 1.25] – small |
| Recovery (a.u.) | 8.92±1.85* | 5.31±1.89 | 0.002 | -1.83 [-2.64; -1.02] – large |
| Thigh temperature (°C) | 35.88±0.73 | 36.22±0.34 | 0.066 | 0.49 [0.08; 0.90] – small |
| Mean radial error (m) | 1.92±0.42* | 2.59±0.67 | 0.025 | 1.50 [0.46; 2.54] – large |
| Bivariate variable error (m) | 1.86±0.59 | 1.79±0.95 | 0.764 | 0.12 [-0.82; 0.58] – trivial |
| Accuracy (m) | 2.76±0.51 | 3.26±1.10 | 0.113 | 0.93 [-0.04; 1.90] – large |
| Average ball speed (m/s) | 28.70±2.30* | 27.80±1.19 | 0.038 | -0.36 [-0.64; -0.09] – small |
| Peak ball speed (m/s) | 29.68±2.64* | 28.51±2.05 | 0.029 | -0.42 [-0.72; -0.12] – small |
| CM<sub>foot</sub> velocity (m/s) | 23.88±6.55 | 24.39±8.27 | 0.224 | 0.07 [-0.21; 0.36] – trivial |
| CM<sub>foot</sub> to knee relative velocity (m/s) | 17.59±3.42 | 18.49±3.89 | 0.274 | 0.24 [-0.14; 0.63] – small |
| Non-preferred hip velocity (m/s) | 10.51±3.19 | 11.41±4.65 | 0.244 | 0.27 [-0.12; 0.65] – small |
| Peak knee angular velocity (rad/s) | 0.53±0.18 | 0.57±0.22 | 0.497 | 0.17 [-0.27; 0.61] – small |
| ROM hip (rad) | 1.30±0.47 | 1.37±0.49 | 0.161 | 0.13 [-0.05; 0.30] – trivial |
| ROM knee (rad) | 1.54±0.46 | 1.57±0.46 | 0.229 | 0.06 [-0.03; 0.14] – trivial |
| Plantarflexion/dorsiflexion at impact (rad) | -0.54±0.47 | -0.54±0.43 | 0.968 | 0.00 [-0.13; 0.14] – trivial |
| Eversion/inversion angle at impact (rad) | -0.07±0.22* | 0.11±0.15 | 0.019 | 0.78 [0.27; 1.30] – moderate |

Note: RPE, ratings of perceived exertion; ROM, range-of-motion; ES, effect size (Cohen's *d*); CL, confidence limits. \*significant difference when comparing Pre- vs Post-RHIR at $p \leq 0.05$ level.

### #Experiment 2

#### Running protocol reliability

Table 2 presents performance indices observed in the running protocol, statistical outcomes from comparisons of these indicators between distinct intervention conditions as well as their concordance. No significant differences were identified for any of the parameters (i.e. MT, WT, BT, TT and DEC) when players performed Control or COOL conditions (*p* = 0.17 to 0.43, *small* effect sizes, -0.34 to 0.38). Running outputs exhibited *moderate*-to-*good* reliability (ICCs = 0.50–0.88; *p* ≤ 0.04) between the two conditions with the exception of DEC (ICC = -0.43; *p* = 0.95). CVs ranged between 2.93% (TT) to maximal 5.87% (BT) while a substantially higher value was observed for DEC (CV = 40.66%).

**Table 2.**
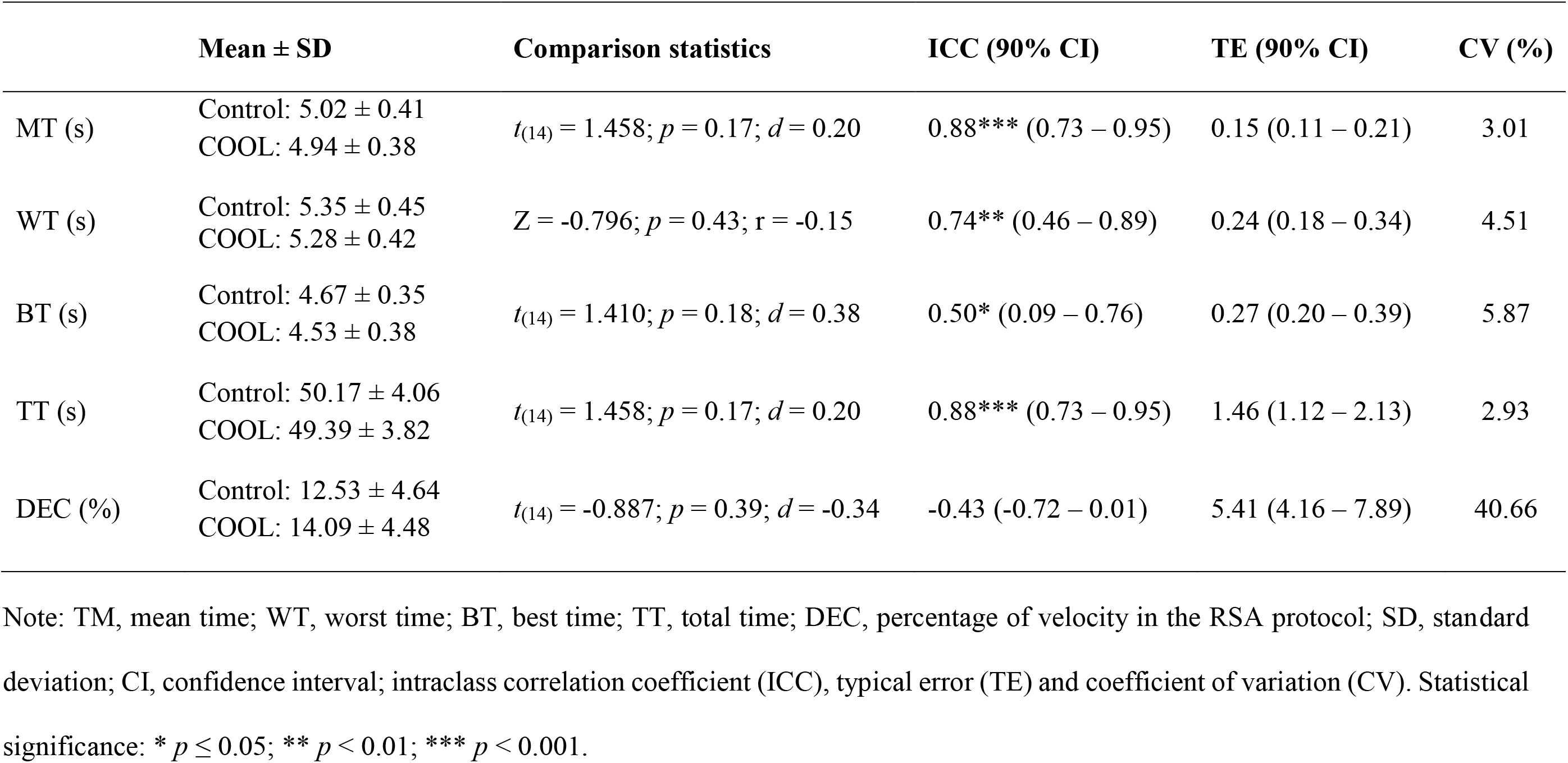
Characterisation of performance indices in the repeated high-intensity running bouts for each condition with reliability measures for responses computed between conditions (n = 15).

#### Recovery treatment effects

Perceptual measures and skin temperature are presented in Figure 1. Responses were similar at baseline (Pre) between the two experimental conditions (*p* = 1.00; r = 0.02 to 0.38). Friedman’s test showed a significant *moderate* main effect for RPE (*X*^2^_(3)_ = 13.408; *p* = 0.004; Kendall’s *W* = 0.30; Figure 1(A)). Pairwise comparisons revealed *largely* increased RPE in the Post-as compared to Pre-Control (+484.85%, 1 a.u. [0; 3]; *p* = 0.02; r = 0.69) whilst RPE was similar across moments in the COOL condition (+75.00%, 0 a.u. [0; 2]; *p* = 1.00; r = 0.35 [*moderate*]). Friedman’s test also detected a *large* main effect for thigh temperature (*X*^2^_(3)_ = 25.711; *p* < 0.001; Kendall’s *W* = 0.57; Figure 1(D)). Pairwise comparisons indicated a *large* decline following COOL as compared to pre-COOL (−8.59%, 3.3 ºC [2.4; 4]; *p* < 0.001; r = 0.88). Thigh temperature was *largely* lower in the COOL as compared to Control at Post moment (−9.09%, 3.2 ºC [2.0; 3.7]; *p* = 0.002; r = 0.88). A main effect in Friedman’s test also occurred regarding the perceived Recovery (*X*^2^_(3)_ = 11.057; *p* = 0.011; Kendall’s *W* = 0.25 [*small*]; Figure 1(C)); however pairwise comparisons lacked statistical significance (e.g. Pre-vs. Post-Control; -23.85%, 1 a.u. [0; 4]; *p* = 0.08; r = -0.65 [*large*]).

**Figure 1.**
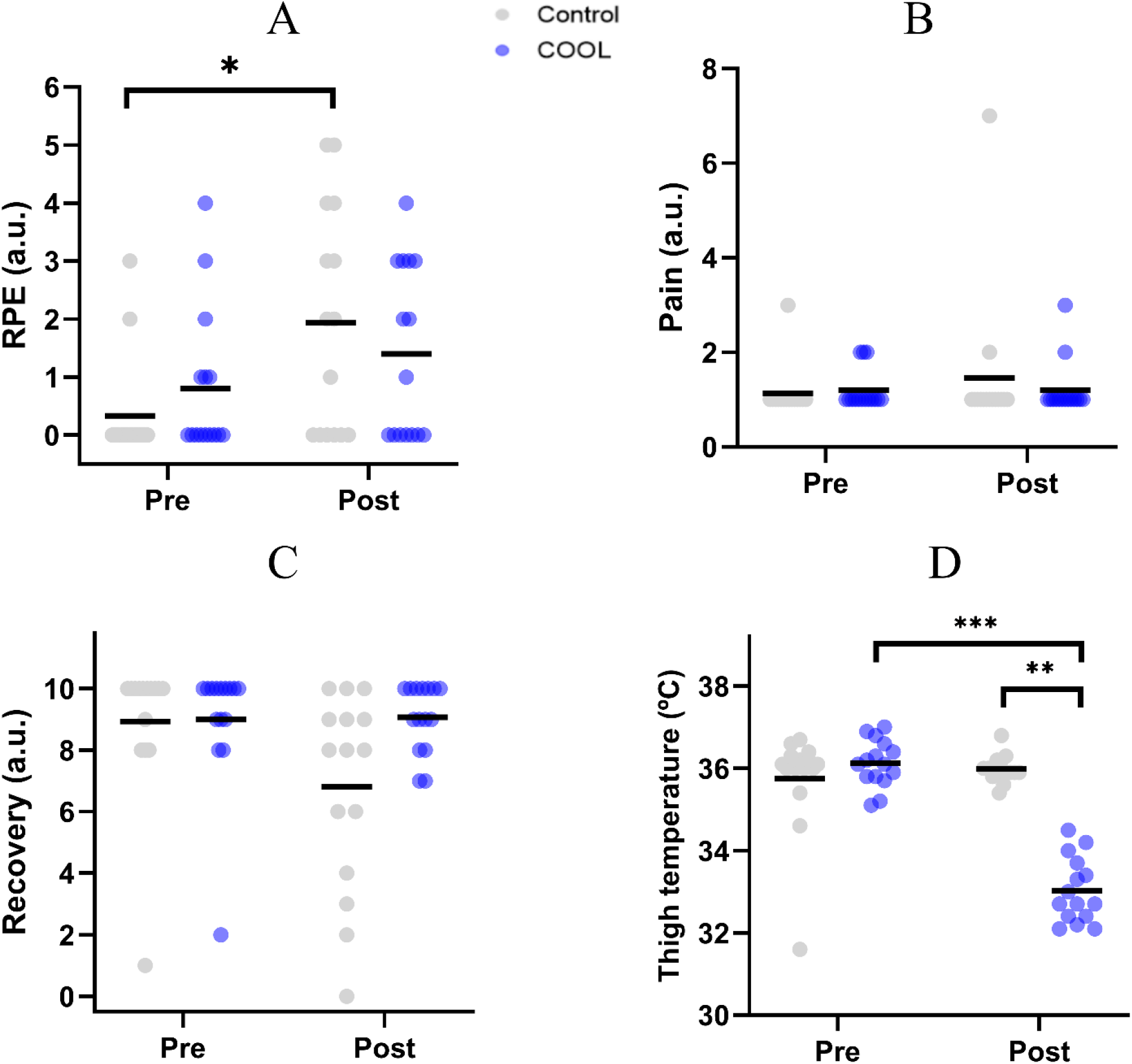
Perceptual measures and skin temperature according to time-moments and conditions. * *p* ≤ 0.05; ** *p* < 0.01; *** *p* < 0.001.

Kicking ball placement-derived indices (mean radial error, bivariate variable error and overall accuracy) and ball speed (average and peak) across the experiment are shown respectively in Figure 2 and Figure 3. These parameters did not differ at baseline when comparing Control and COOL conditions (*p* = 0.39–0.78; *d* = 0.11–0.36 and *p* = 0.47–0.78; *d* = 0.10–0.29 respectively). There was a *large* main Time × Condition interactive effect of borderline significance concerning mean radial error (*F*_(1, 14)_ = 4.286; *p* = 0.057; η^2^ = 0.234). Pairwise comparisons indicated a *large* significant increase in mean radial error after Control condition as compared to baseline (+50.26%, 0.97 m [0.40; 1.54]; *p* = 0.003; *d* = -1.12) while no significant pre-post variations existed in COOL intervention (+12.44%, 0.24 m [-0.43; 0.91]; *p* = 0.45; *d* = -0.25 [*small*]).

**Figure 2.**
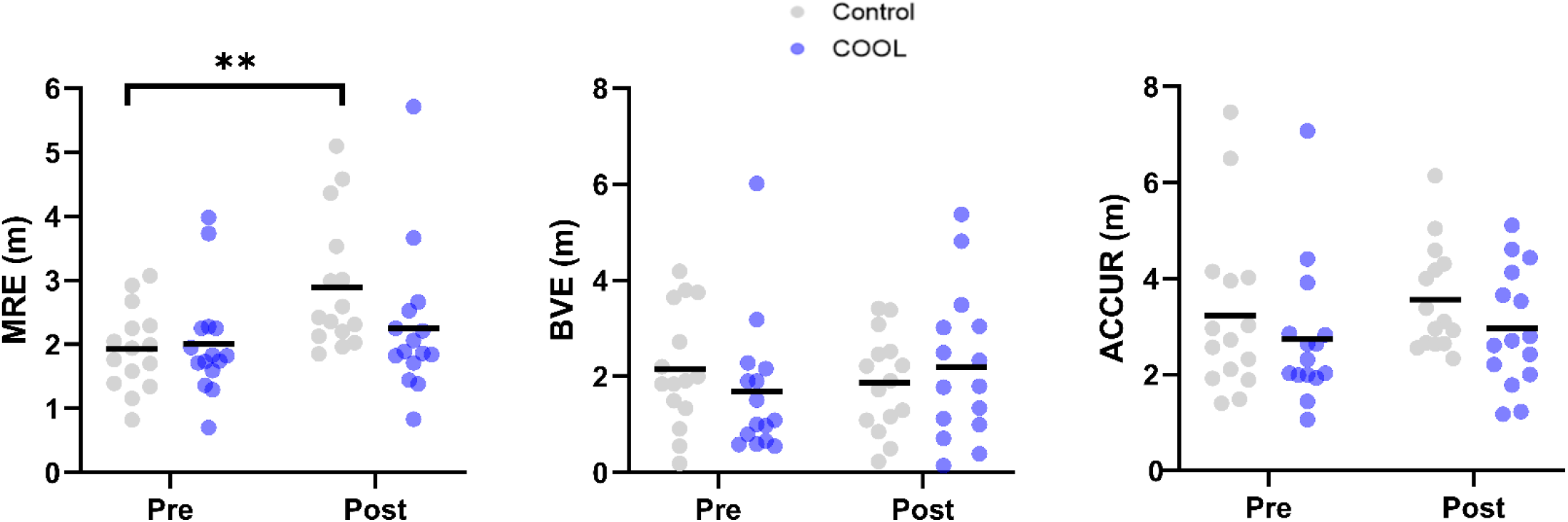
Ball placement-derived indices according to time-moments and conditions. ** *p* < 0.01.

**Figure 3.**
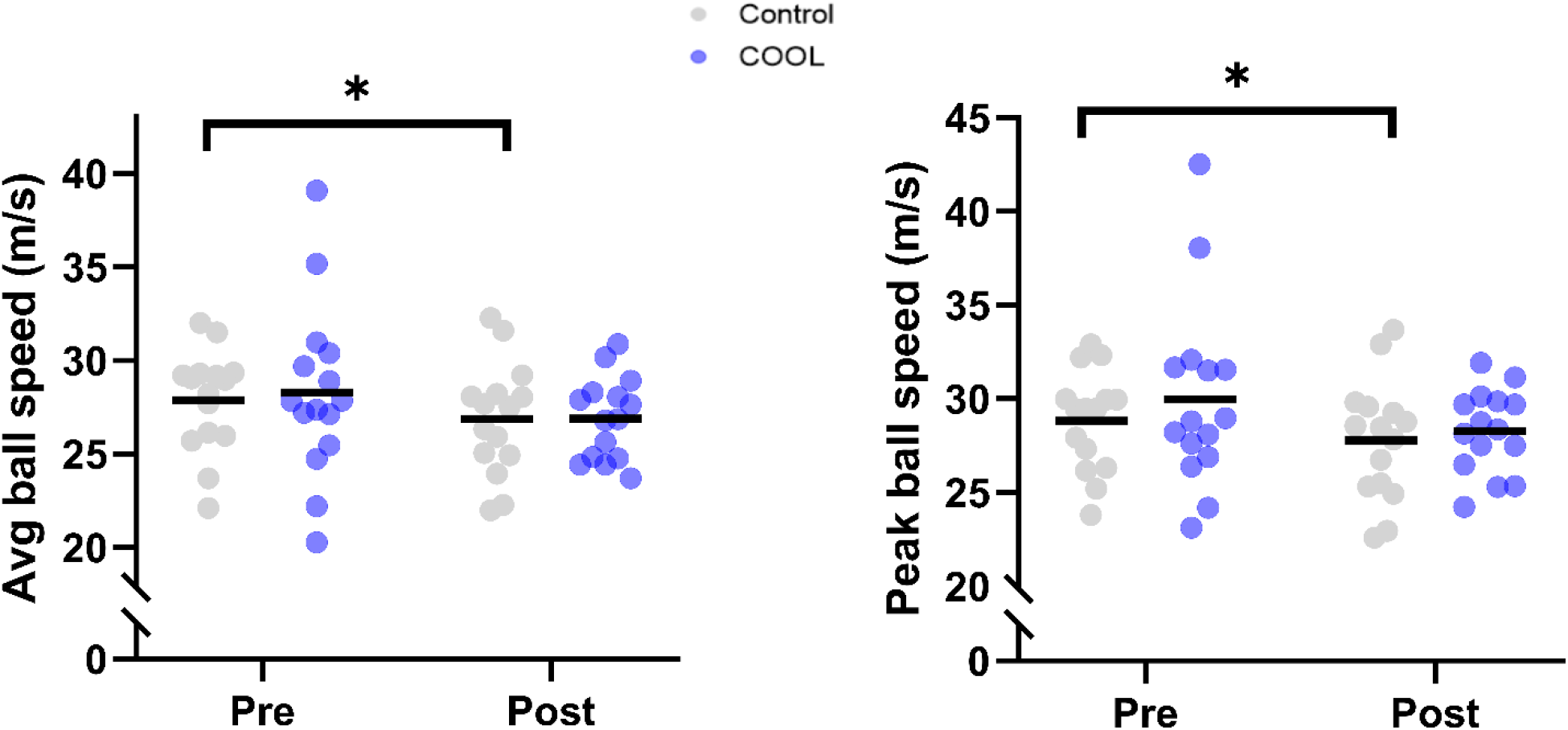
Ball speed indices [average (Avg) and maximal (peak)] according to time-moments and conditions. * *p* ≤ 0.05; ** *p* < 0.01.

No significant *small* Time × Condition effects were found for ball speed (average: *F*_(1, 14)_ = 0.126; *p* = 0.73; η^2^ = 0.009; peak: *F*_(1, 14)_ = 0.394; *p* = 0.54; η^2^ = 0.027). Separate *large* Time effects were significant for both average (*F*_(1, 14)_ = 6.649; *p* = 0.02; η^2^ = 0.322) and peak ball speed (*F*_(1, 14)_ = 6.580; *p* = 0.02; η^2^ = 0.322). Pairwise comparisons revealed significant *small* declines (both *d* = 0.35) in ball speed following the Control condition (average: -3.62%, -1.01 m/s [-1.95; -0.06]; *p* = 0.04 and peak: -3.57%, -1.04 m/s [-1.89; -0.18]; *p* = 0.02) but in COOL condition (average: -4.92%, -1.40 m/s [-3.38; 0.59]; *p* = 0.15; *d* = 0.38 and peak: -5.67%, -1.70 m/s [-3.83; 0.42]; *p* = 0.11; *d* = 0.44).

Regarding the movement kinematics (Figure 4), in general the measures also showed no between-condition differences at baseline (*p* = 0.14–0.84; *d* = 0.02–0.42). One exception occurred for the value of ankle eversion angle at impact (main Time × Condition effect: *F*_(1, 14)_ = 5.339; *p* = 0.04; η^2^ = 0.276 [*large*]), which had a significant pairwise difference among conditions in the Pre (355.56%, -0.32 rad [-0.58; -0.06]; *p* = 0.02; *d* = 1.02 [*large*]) but this was not the case in the Post-intervention (33.33%, 0.01 rad [-0.26; 0.29]; *p* = 0.91; *d* = -0.06 [*trivial*]; Figure 4(H)). Finally, there was a *large* main Time × Condition interactive effect in reference to CM_foot_ velocity (*F*_(1, 14)_ = 6.538; *p* = 0.02; η^2^ = 0.318). In particular CM_foot_ velocity was *moderately* faster in the COOL as compared to Control in the Post-intervention moment (+21.85%, 4.61 m/s [0.14; 9.08]; *p* = 0.04; *d* = 0.60; Figure 4(A)).

**Figure 4.**
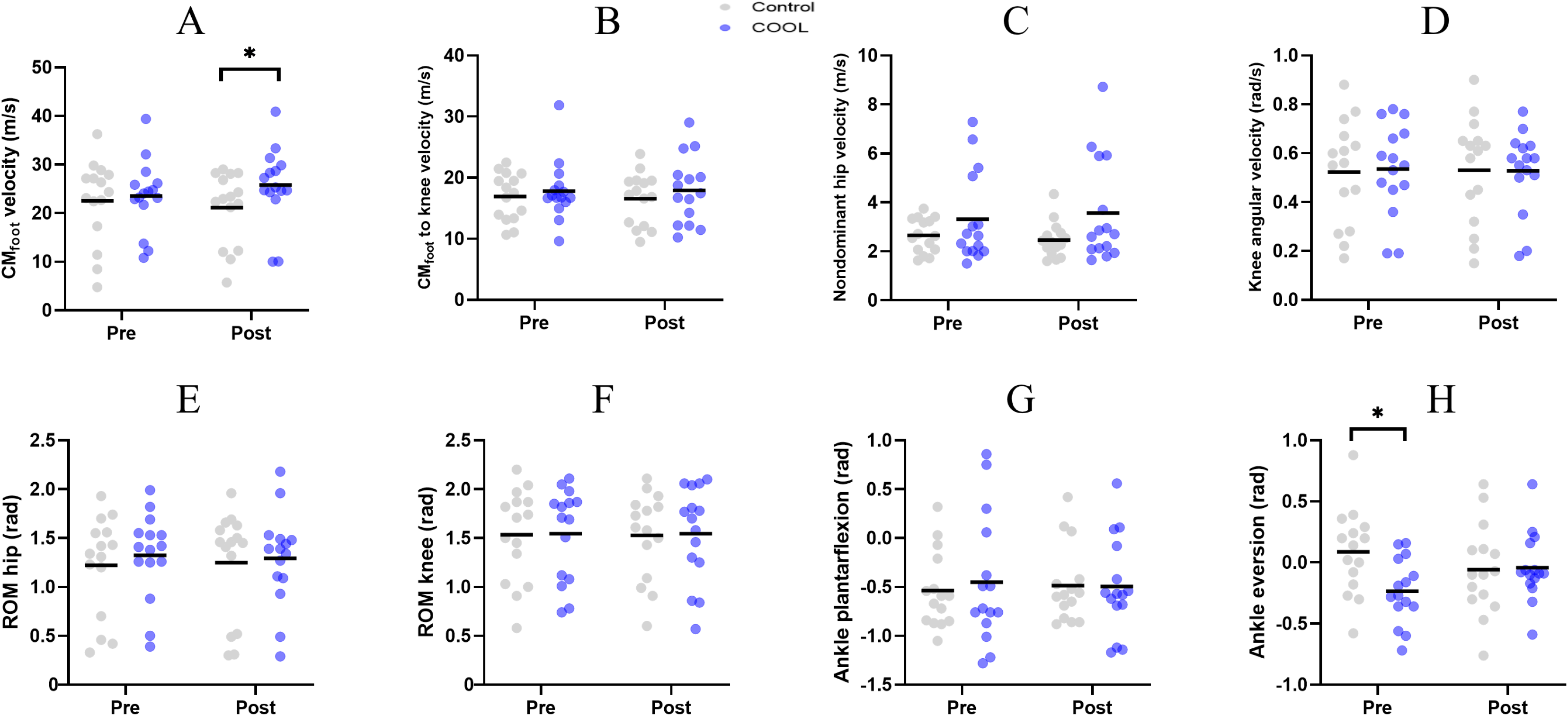
Kinematics of the lower contact limb according to time-moments and conditions. * *p* ≤ 0.05.

## DISCUSSION

The main purposes of the present investigation were to investigate, in youth soccer players and under an environmental stressor (hot temperature) condition (i) whether repeated high-intensity running (RHIR) efforts immediately modify subsequent kinematics and performance (ball placement and velocity) outputs in kick attempts performed from the edge of penalty area and (ii) the effectiveness of applying a brief local cooling pack (COOL) on the thigh as a potential recovery intervention following this type of locomotor exercise and its consequences on kicking measures. Our first hypothesis was rejected since the intense exercise provoked negative changes notably in ball placement aspects of kicking ability (inducing kicks generally farther from the target in goalpost upper corners) while ball speed was affected to a small extent. Conversely and partly in line with our second hypothesis, the use of 5-minutes COOL following RHIR bouts promoted benefits in perceptual measures (internal load), kinematics (CM_foot_) and performance (both ball placement and velocity indicators) of soccer kick attempts as compared to the control condition. Hereinafter, we discuss the transient negative impact of RHIR exercise and the ergogenic effects provided by COOL during the acute recovery phase, emphasizing the distinct responses of kicking components to both exercise/cool-down.

In general, RHIR bouts impaired both kinematics and performance components of soccer kicking in youth under-17 players. These include distal mechanics of contact limb as illustrated by modification of ankle eversion to inversion pattern at foot-ball impact moment. In addition, mean radial error substantially increased following the running protocol potentially performance. Recent related research has notably shown that greater ankle inversion is associated to higher mean radial error in kick attempts from entrance of penalty area (Palucci Vieira et al., 2022a). Ball speed verified in the present experiment was within the range (22–32 m/s) of age-matched players (Nunome et al., 2006; Vieira et al., 2018) while the mean radial error was slightly greater than in penalty trials or 15-m kicks (0.90–1.50 m; (Russell et al., 2011; Vieira et al., 2018)). According to a recent review on the topic (Palucci Vieira et al., 2021b) only two studies assessing the acute impact of RHIR mode of exercise on subsequent kicking aspects of youth soccer currently exist. Non-significant (trivial-to-large) exercise effects on accuracy were previously verified but no concomitantly information on kicking velocity was provided (Gharbi et al., 2017; Masmoudi et al., 2016). Of note, these works were conducted in players mean aged 14.6 years-old, that certainly fell in the circum-PHV stage (Buchheit and Mendez-Villanueva, 2013) while the present players were all post-PHV. More mature players perform faster bouts than developing pre/circum-PHV peers in repeated sprints of equalized volume (Selmi et al., 2020). Consequently, the impaired ball placement ability following RHIR efforts verified in the current investigation in youth aged under-17 but not in under-15s in literature can be potentially attributed to a superior disturbance in homeostasis experienced in the former due to their higher intense locomotor output profiles. Notwithstanding, kick task constraints as attempts performed 6.1 m apart from a small-dimension (2.44 × 1.22 m) goal/target aiming only its centre (Gharbi et al., 2017; Masmoudi et al., 2016) prevents such a direct comparison to our results or otherwise could question whether prior evidence of no changes on ball placement ability in youth soccer following RHIR is due to low-challenging kick testing demands. Our findings are in accordance with those of Rampinini et al. (2008) who verified impaired short passing performance in youth players following RHIR that attempted to reproduce most demanding phase of matches (10 × 40 m with one change-of-direction). Importantly, exercising with RHIR in addition to a heat condition are recognised to cause central neurotransmission deficits and a drop in muscle activity (Goodall et al., 2015; Meeusen et al., 2006; Perrey et al., 2010), both determinant aspects to proficiency in targeting the ball when kicking (Palucci Vieira et al., 2022a; Palucci Vieira et al., 2021b). To summarize, intense and consecutive running bouts (e.g. match-play worst case scenarios) could impair soccer kicking in youth under-17 age-group, mainly the ability of players placing the ball in the goalpost upper corners.

It is necessary to highlight a discrepant behaviour of kicking velocity and ball placement responses to the exercise whilst recovery intervention effect was similar for both aspects. In fact central and peripheral signalling paths/measures responsible for kick accuracy and velocity are not the same (Palucci Vieira et al., 2022a), which a priori help justify differences. However, exercise-related fatigue affects to a similar extent the functioning of brain regions/waves determinants for ball speed (frontal theta) and placement (occipital alpha) (Baumeister et al., 2012) meaning that the problem could be more at limb level. Evidence that vastus lateralis RMS may be unchanged as a function of repeated sprints execution and have fast recovery has been provided (Billaut and Basset, 2007). EMG amplitude indices such as RMS and integral were recently demonstrated to affect ball speed in teenagers (unpublished observations from our laboratory). Also, biceps femoris activity that is related to ball placement in youth soccer seems more consistently modified by repeated intense bouts across studies (Hautier et al., 2000; Timmins et al., 2014; Zarrouk et al., 2012) given the high strain on the hamstrings during decelerations separating RHIR efforts. On the other hand, there is a strong linear relation amongst kicking ball speed and chance of goal attempt to become blocked (Palucci Vieira et al., 2021b). Whether players intrinsically adopted a possible strategy of trying to maintain the velocity output under fatigue, with impaired control (e.g. ankle joint) this may have resulted in worst ball placement following RHIR.

Aside from a worst ball placement induced by RHIR, it is necessary to highlight that reductions in ball speed due to this mode of exercise although significant were small and does not surpass minimal detectable difference (∼1.27 m/s; Palucci Vieira et al. (2022b)). Even though such little reductions caused by separate RHIR efforts, official competition demands induced moderate-to-large declines in post-match ball speed elsewhere in youth from all playing positions (Izquierdo et al., 2020). This reinforces the potential role of COOL in practice to prevent this picture given the ergogenic effects it had on foot and ball velocities according to our analysis. Notwithstanding, while technological or logistic difficult still exist limiting collection of advanced kicking technique features during real-world events, simulated game-play running demands (e.g. RHIR efforts) seems a pertinent strategy (Palucci Vieira et al., 2021b; Rampinini et al., 2008; Sánchez-Sánchez et al., 2014). In this sense, according to existing reviews (Lopes-Silva et al., 2019; Paul and Nassis, 2015), the day-to-day repeatability of the specific on-field RHIR protocol used has not been determined before this experiment; a single study with similar sprint number/larger course (10 × 40 m; no mention to environmental condition) including youth elite European soccer players age-matched to our sample reported only average bout duration as a reference parameter (r = 0.94), such that it do not always reflect various components of repeated sprints performance (e.g. lack of average-best bouts association). Here the 10 × 30 m RHIR model showed consistent outcomes between distinct experimental conditions in the heat also in peak, worst and accumulated effort durations in addition to no between-day statistically significant differences in all measures (indicative of low random and systematic bias, respectively), thereby providing preliminary evidence at first time supporting global reproducibility of a specific tool to South American academy players.

One key finding of the present analysis was that a cooling intervention using the application of ice pack on the quadriceps/hamstrings across following RHIR performed in the heat reduced the perception of effort while seemingly preventing negative consequences such as worst ball placement, CM_foot_ and ball speed declines as observed in the control condition–passive resting during a time-matched period–but not in the COOL condition. There was a trend represented by non-significant (*p* = 0.08) large-sized decrease in perceived recovery during post-control while this was not the case in COOL condition. Among the mechanisms possible acting, a forceful reduction in local temperature can have counteracted declines in neuromuscular output observed under heat (Matsuura et al., 2015). Another aspect to appraise these results is that a 5-min induced COOL, despite promoting substantial decreases in skin (Figure 1) and subcutaneous temperatures (Myrer et al., 1997), it can cause limited decline in intramuscular temperature as compared to the rested state (e.g. ∼0.64ºC; Zemke et al. (1998)). This is important owing to the strong relationship between declines in muscle temperature and lower limb (sprint) performance (Mohr et al., 2004). Locomotor outputs have been indirect markers related to kicking quality in youth (Sporiš et al., 2007), meaning that likely exist shared mechanisms responsible by those explosive actions and if it hold true, an exacerbated muscle temperature drop may lead also into kick deficits. In a meta-analysis, the effectiveness of a commonly used COOL mode namely cold-water immersion was reported to similarly alleviate RPE but had no meaningful effects in power performance in teenagers (Murray and Cardinale, 2015). Also, long periods (e.g. 2 × 15 min) of ice pack maintenance following interval sprints session are recognised to cause bionegative adaptations such as decreased anabolic response in youth team sport athletes (Nemet et al., 2009). Taken together to our results, the premise that COOL has a time-dependent effect on ensuing performance (Bleakley et al., 2012; Fischer et al., 2009; Peiffer et al., 2009) seems well supported as a brief cooling intervention was effective in some instances–especially concerning ball placement–or at least not damaged kicking outputs whilst attenuated exercise effort and recovery perceptions. Thus, we provide evidence for the first time that a short-term local ice pack application following RHIR may assist players in produce subsequent kicks with a greater likelihood of success than resting passively, ameliorating exercise-induced fatigue consequences in skilled performance in a youth soccer context.

Innovative characteristics of the present investigation include empirical testing of whether COOL has benefits to recover (intense)exercise-induced kicking performance loss under heat stress, which fills an important knowledge gap with potential to modify current practices (Bleakley et al., 2012; Palucci Vieira et al., 2021b). Conversely, there are various caveats that should be made which collectively may limits generalizability of this study findings. We have expected that RHIR would promote an acute increase pain sensation while COOL act in its recovery and this was not confirmed since the post-exercise perception of pain was similar to baseline levels. The lack of an extended familiarization of players with such monitoring tool is arguably one of the reasons or otherwise it reflects only a general state of pain and thus scales that compute local muscle soreness are recommended. Advanced measures of body temperature (e.g. infrared thermography) were lacking making unclear the actual physiological impact of COOL treatment over whole working muscles group. When designing the task, we opted to avoid opposition players contesting kicks in an attempt to reduce potential undesirable inter-trial/condition variability interfering with the treatment effects. Despite increasing experimental control, this approach greatly reduces the external validity. Finally, it is yet to be determined the extent to which findings observed in the present investigations are transferable to other intervention format (e.g. cooling chamber), task conditions such as penalty kicks and crowded competition environments.

## CONCLUSIONS

Repeated high-intensity running bouts acutely affect soccer kicking performance in youth academy players. Ankle kinematic adjustments at foot-ball impact instant and consequent ball placement are notably impaired following this mode of exercise while kicking velocity aspects are impacted to a small extent. A protocol consisting in 10 × 30 m running efforts interspersed by 30 s intervals is a reliable method in a hot environment, consistent in day-to-day basis. Rather than providing a passive resting period, including 5 minutes of two ice packs applied respectively to the quadriceps and hamstrings muscles of contact limb may favour recovery of wellbeing aspects and overall soccer kicking parameters. Thus, short-term cryotherapy plays an important role in counteracting fatigue-related declines experienced by youth under-17 players in terms of kicking outputs following repeated high-intensity running efforts in the heat.

## Data Availability

All data produced in the present work are contained in the manuscript

https://osf.io/bd8zv/

## DATA AVAILABILITY STATEMENT

The raw data supporting the conclusions of the current manuscript has been made publicly available at https://osf.io/bd8zv/ (Open Science Framework – OSF).

